# Contact tracing efficiency, transmission heterogeneity, and accelerating COVID-19 epidemics

**DOI:** 10.1101/2020.09.04.20188631

**Authors:** Billy J. Gardner, A. Marm Kilpatrick

## Abstract

Simultaneously controlling COVID-19 epidemics and limiting economic and societal impacts presents a difficult challenge, especially with limited public health budgets. Testing, contact tracing, and isolating/quarantining is a key strategy that has been used to reduce transmission of SARS-CoV-2, the virus that causes COVID-19. However, manual contact tracing is a time-consuming process and as case numbers increase it takes longer to reach each cases’ contacts, leading to additional virus spread. Delays between symptom onset and being tested (and receiving results), and a low fraction of symptomatic cases being tested and traced can also reduce the impact of contact tracing on transmission. We examined the relationship between cases and delays and the pathogen reproductive number R_t_, and the implications for infection dynamics using a stochastic compartment model of SARS-CoV-2. We found that R_t_ increases sigmoidally with the number of cases due to decreasing contact tracing efficacy. This relationship results in accelerating epidemics because R_t_ increases, rather than declines, as infections increase. Shifting contact tracers from locations with high and low case burdens relative to capacity to locations with intermediate case burdens maximizes their impact in reducing R_t_ (but minimizing total infections is more complicated). Contact tracing efficacy also decreased with increasing delays between symptom onset and tracing and with lower fraction of symptomatic infections being tested. Finally, testing and tracing reductions in R_t_ can sometimes greatly delay epidemics due to the highly heterogeneous transmission dynamics of SARS-CoV-2. These results demonstrate the importance of having an expandable or mobile team of contact tracers that can be used to control surges in cases, and the value of easy access, high testing capacity and rapid turn-around of testing results, as well as outreach efforts to encourage symptomatic infections to be tested immediately after symptom onset.

**Author Summary:** A key tool in the control of infectious diseases is contact tracing – the identification of individuals who have contacted the case and may have been infected by a newly detected case. However, to successfully contact and quarantine individuals requires time, and as cases rise, this can result in delays in reaching contacts during which time they may infect other people. Here we examine the quantitative relationships between increasing case numbers, contact tracing efficiency, and the pathogen reproductive number R_t_ (the number of cases infected by each case) and how these relationships vary with delays and incomplete participation in the testing and tracing process. We built

## Introduction

Severe acute respiratory syndrome coronavirus 2 (SARS-CoV-2) emerged in late 2019 spread globally in early 2020 and resulted in rapidly growing local epidemics, large scale mortality, and strains on hospital capacity in many countries (1-4). Initial outbreaks in most countries and US states were arrested only by severe control measures including closing all but essential businesses as well as schools, churches, and other organizations (5), while a few countries were able to limit transmission, at least temporarily, primarily with public health measures (6-8). Severe disease control measures have had devastating impacts on economies and societies (5). Most countries and US states are now attempting to re-open as many business sectors and activities as possible while avoiding a rapid rise in infections.

Although self-isolation, social distancing, and mask wearing have reduced the transmission of SARS-CoV-2, additional interventions, including business closures and working from home, have often been required to keep the pathogen reproductive number Rt below 1 (5, 9, 10), especially in the United States. One public health strategy that has been used effectively to limit transmission in some countries is testing symptomatic individuals, tracing their contacts to people they may have infected, and isolating infected individuals and quarantining people that may have become infected but have yet to show symptoms or test positive for the virus (hereafter abbreviated T-CT-I/Q) (7, 10, 11). If contacts of cases can be found and quarantined or isolated before or during their infectious period, this can limit onward spread of the virus.

Numerous studies have examined the effectiveness and limitations of T-CT-I/Q on transmission of SARS-CoV-2 (11-18). Many studies have shown that T-CT-I/Q can substantially reduce the pathogen reproductive ratio, R_t_, but its efficacy depends on the importance of pre-symptomatic and asymptomatic transmission, delays between symptom onset and being tested, and the fraction of infections that are tested and traced (11, 15, 18). Previous studies have explored various parameter values for contact tracing efficacy by varying the fraction isolated, the fraction symptomatic, and the contribution to transmission of undetected infections (11, 15, 18). A key unexplored challenge in implementing T-CT-I/Q is that tracing contacts and ensuring they can safely quarantine or isolate is a time-consuming process which results in delays between detecting a case and successfully quarantining their contacts. This delay reduces the effectiveness of contact tracing as cases increase. Previous studies have assumed fixed values for contact tracing parameters and thus have not addressed this issue.

Our aim was to examine the relationship between increasing cases, contact tracing efficacy, and the pathogen reproductive ratio, R_t_, and to examine the potential outcomes for disease dynamics. We built a compartment model of SARS-CoV-2 transmission, parameterized it with data from the literature, and examined how R_t_ varied with number cases traced, delays between symptom onset and the start of contact tracing, the numbers of contacts per case, and different fractions of symptomatic cases being tested and traced. We also simulated a stochastic version of the model with and without contact tracing to examine how reductions in Rt affected variation in the timing of epidemics.

## Methods

We built a susceptible-exposed-infected-recovered (SEIR) compartment model of SARS-CoV-2 that included four compartments for infected individuals that reflect symptom severity (asymptomatic, *I_a_*, pre-symptomatic, *I_ps_*, mildly symptomatic, *I_ms_*, and severely symptomatic, *I_ss_*) (Fig S1). The equations of the model are:

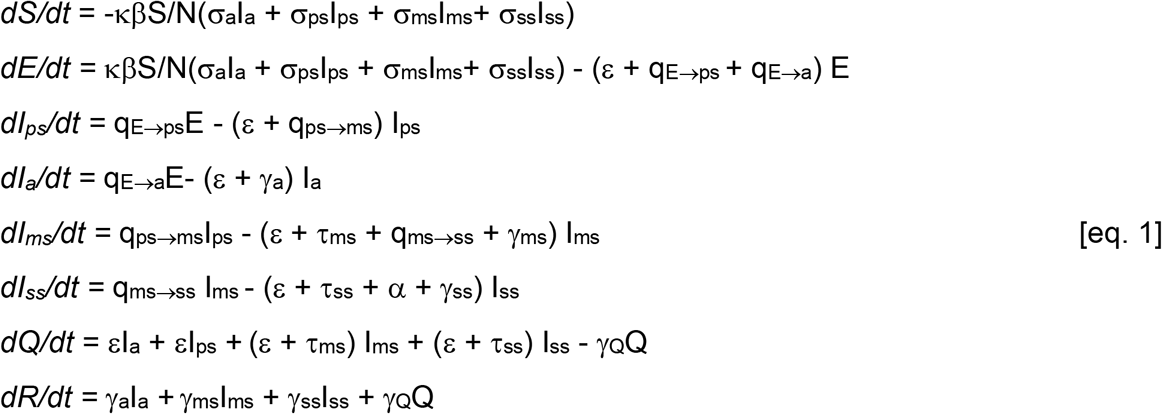

where κ is a social distancing factor between 0 and 1 that scales the contact rate β, σ are relative infectiousness values for each of the I classes, q are transition rates between classes given by the subscripts separated by the arrow (q_E→ps_ is the transition rate between the E and I_ps_ classes), ε is the rate of removal by contact tracing from the E or I classes to the quarantined class Q, τ are the removal rate by testing of mildly symptomatic or severely symptomatic infected individuals, α is the disease-caused death rate, and γ are the recovery rates to the R class.

The contact tracing removal rate ε is given by:

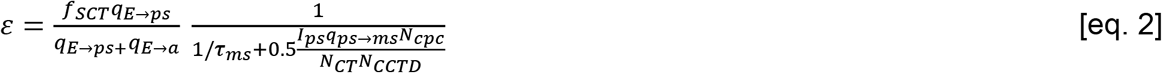

The first term is the fraction of infections traced; f_SCT_ is the fraction of symptomatic infections that are traced of those detected with test removal rate τ_ms_ before they recover or progress to severe symptoms; f_SCT_ is equal to f_tmstr_*τ_ms_/(τ_ms_+q_ms→ss_+γ_ms_); f_SCT_ is multiplied by the ratio of transition rates [q_E→ps_/(q_E→ps_+q_E→a_)] which is the fraction of infections that are symptomatic; ftmstr is the fraction of mildly symptomatic cases detected by testing that were traced. We didn’t include tracing from severely symptomatic cases because, by the time an infection progresses to severe symptoms 5-8 days after symptom onset (19, 20), their contacts will already have finished most of their infectious period, and quarantining their contacts will have little effect.

The second term is the inverse of the time between symptom onset of the symptomatic infection and being removed by contact tracing which is the sum of the delay between symptom onset and receiving a test result, 1/τ_ms_, and the average time needed to reach the contacts of the newly detected cases (which is half the total time to call all contacts). q_ps→ms_I_ps_ is the rate of new symptomatic infections that need to be traced, N_cpc_ is the average number of contacts per case, N_CT_ is the number of contact tracers, and N_CCTD_ is the number of calls each contact tracer can make each day.

We parameterized the model with data from the literature, with all rates in days^-1^ (Table 1) and used the baseline contact tracing capacity standards suggested by the US National Association of County and City Health Officials (15 contact tracers per 100,000 people; https://www.naccho.org/uploads/full-width-images/Contact-Tracing-Statement-4-16-2020.pdf). We note that while notifying individuals that they have had contact with a case can be done quickly (especially with using a cell-phone tracing app; (18)), successfully ensuring a contact has their needs met (including food, medicine, clothing) to quarantine in a safe space where they won’t infect their household members requires substantially more time (https://www.cdc.gov/coronavirus/2019-ncov/php/notification-of-exposure.html). We assumed the approximate duration required for a successful contact tracing call was 40 min, resulting in N_CCTD_ = 12

**Table 1.** Parameter values for the model. All rates are in days^-1^ and many are based on the inverse of measured durations.

| Parameter | Value or range | Description | Reference or Derivation |
| --- | --- | --- | --- |
| $\tau_{ms}$ | 0.2-1 | Testing removal rate for $I_{ms}$ (1/delay from onset to testing & tracing) | Scenarios explored |
| $\beta$ | 0.37 | Contact rate | Set to give plausible pre-lockdown $R_0 \cong 2-3$ (29) |
| $f_{tmstr}$ | 0.5 or 1 | Fraction of mildly symptomatic cases detected by testing whose contacts were traced | Scenarios explored |
| $\kappa$ | 0-1 | Social distancing factor | Adjusted to produce $R_t \cong 1.2-1.7$; consistent with data post-lockdown; (29) |
| $\alpha$ | 0.0025 | Disease caused death rate | estimated from IFR* $(\gamma_{ms} + \gamma_{ss} + q_{E \rightarrow a}) / (1 - IFR) =$ using infection fatality ratio IFR = 0.0066; (30, 31) |
| $q_{E \rightarrow a}$ | 0.078 | Transition rate exposed to asymptotically infected | Estimated from $q_{E \rightarrow a} = f_{asympt}(q_{E \rightarrow ps}) / (1 - f_{asympt})$ using fraction asymptomatic ($f_{asympt} = 0.2$); (32) |
| $q_{E \rightarrow ps}$ | 1/3.2 | 1/(duration latent period) | (28) |
| $q_{ps \rightarrow ms}$ | 1/2.3 | 1/(duration pre-symptomatic period) | (28) |
| $q_{ms \rightarrow ss}$ | 1/8 | 1/duration mild symptoms | (19, 20) |
| $\gamma_a$ | 1/8 | Asymptomatic recovery rate | Based on average ratio of shedding durations compared to symptomatic cases (33) |
| $\gamma_{ms}$ | 1/5, | Mild infection recovery rate | (28) |
| $\gamma_{ss}$ | 1/10.7 | Severe symptom recovery rate | (20) |
| $\gamma_Q$ | 1/14 | Quarantined recover rate | Does not affect dynamics |
| $\sigma_a$ | 1 | Relative infectiousness asymptomatic:mildly<br>symptomatic | (33) |
| $\sigma_{ps}$ | 1.81 | Relative<br>infectiousness pre-symptomatic:mildly<br>symptomatic | (28) |
| $\sigma_{ms}$ | 1 | Relative<br>infectiousness | (reference level) |
| $\sigma_{ss}$ | 0.008 | Relative<br>infectiousness severe<br>symptoms:mildly<br>symptomatic | (28) |

We used the next generation matrix technique to derive an expression for the pathogen reproductive ratio R_t_ (21):

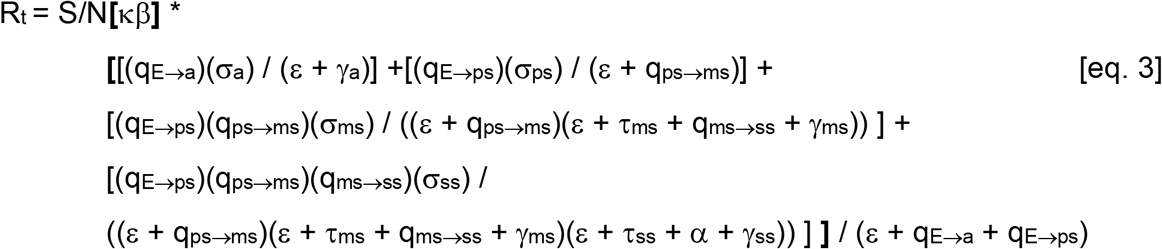

This expression can be understood as the fraction of the population that is susceptible, S/N multiplied by the contact rate β (which is scaled by the social distancing factor κ), multiplied by the sum of four terms: one for each infected class. Each of the four terms includes the product of the transition rates q to reach that class from the E class multiplied by the infectiousness of that class, σ, divided by the recovery, testing and tracing rates leaving that I class and the I classes before it. Finally, all four terms are divided by the rates hosts leave the E class which normalizes the rates moving along each infection pathway.

We examined how R_t_ and R_0_ (R_t_ = R_0_ at beginning of epidemic when S ≅ N) varied with different numbers of new symptomatic cases detected (a fraction of the newly symptomatic infection, *I_ps_* q_ps→ms_ in eq. 2), delays between symptom onset and the start of contact tracing (1/τ_ms_ in eq. 2), and numbers of contacts per case (N_cpc_ in eq. 2). Note that because these three quantities occur as a ratio (with the number of contact tracers) in equation (2), any combination of values that produce the same number of case-contacts per contact tracer calls per day will produce the same value of R_t_. We illustrated the effect of decreasing contact tracing efficiency as infections increased on disease dynamics and R_t_ by simulating a deterministic version of the model in eq. 1 and plotted R_t_ in real time as an epidemic swept through a population over one year. We also performed a simple sensitivity analysis by determining how much R_t_ varied with a ten percent increase or decrease in each model parameter (Fig S2).

Finally, we explored the implications of stochastic variability and contact tracing on infection dynamics in a scenario roughly based on a moderate size city with partly effective social distancing. We simulated a stochastic version of the model given by eq 1 where the number of new infections was drawn from a negative binomial distribution with mean equal to R_t_ and dispersion parameter 0.16 which is intermediate between available estimates for COVID-19 (22-24). We modeled a scenario of a population of 100,000 people where non-pharmaceutical interventions reduced contact rates by 30% (κ=0.7) resulting in R_0_ with/without contact tracing of 1.32/1.67, testing and contact tracing took place after an average of 1/τ_ms_ = 5 days after symptom onset, and half of symptomatic cases detected by testing being traced (f_tmstr_ = 0.5). We examined different initial numbers of latently infected individuals, E, at the start of the epidemic to understand how stochastic variation could impact the timing of epidemics when initial infections were low.

R code to reproduce the results is available from: https://github.com/marmkilpatrick/Contact-Tracing-Efficiency

## Results

The effectiveness of contact tracing in reducing the pathogen reproductive number, R_t_, was highly dependent on three factors: the number of cases being traced (given a fixed number of contact tracers), the delay between symptom onset and the start of tracing, 1/τ_ms_, (including getting tested and receiving result), and the fraction of symptomatic cases that get traced (Figs 1-3).

**Figure 1.**
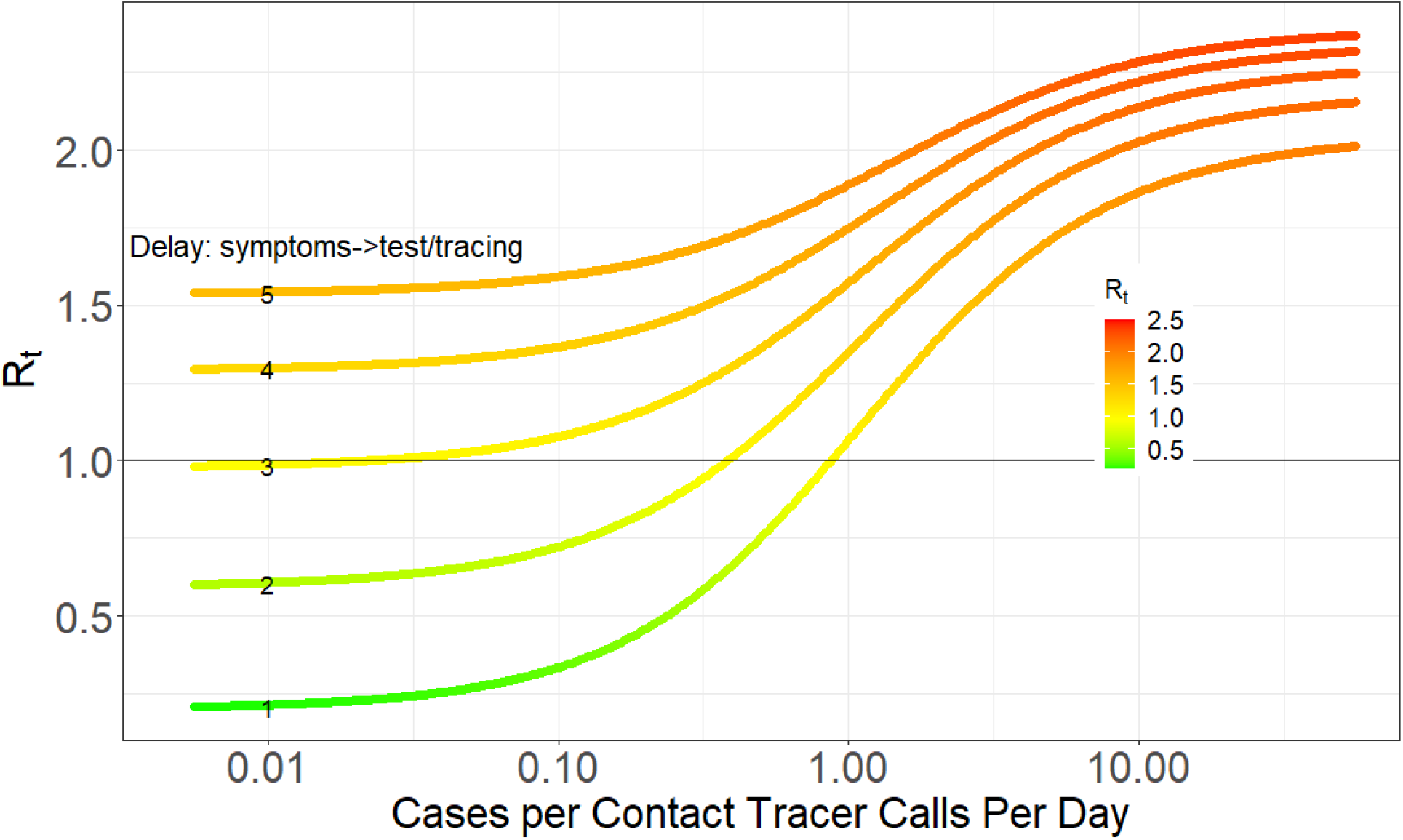
**Pathogen reproductive ratio R_t_ plotted against the number of new cases per contact tracer calls per day for 1-5 day delays (**1/τ_ms_**) between symptom onset and the start of contact tracing (including getting tested and receiving result), 10 contacts per case (so the average number of contacts each tracer has to reach each day is 10x the x-axis values). With no contact tracing R_t_ increases from 2.03 to 2.37 as the delay** 1/τ_ms_ **increases from 1 to 5 days, which is evident in the y-axis difference between curves in the upper right of the graph where new case burdens are so high contact tracing is ineffective. Delays are indicated by the small numbers on each curve in the left of the plot**.

**Figure 2.**
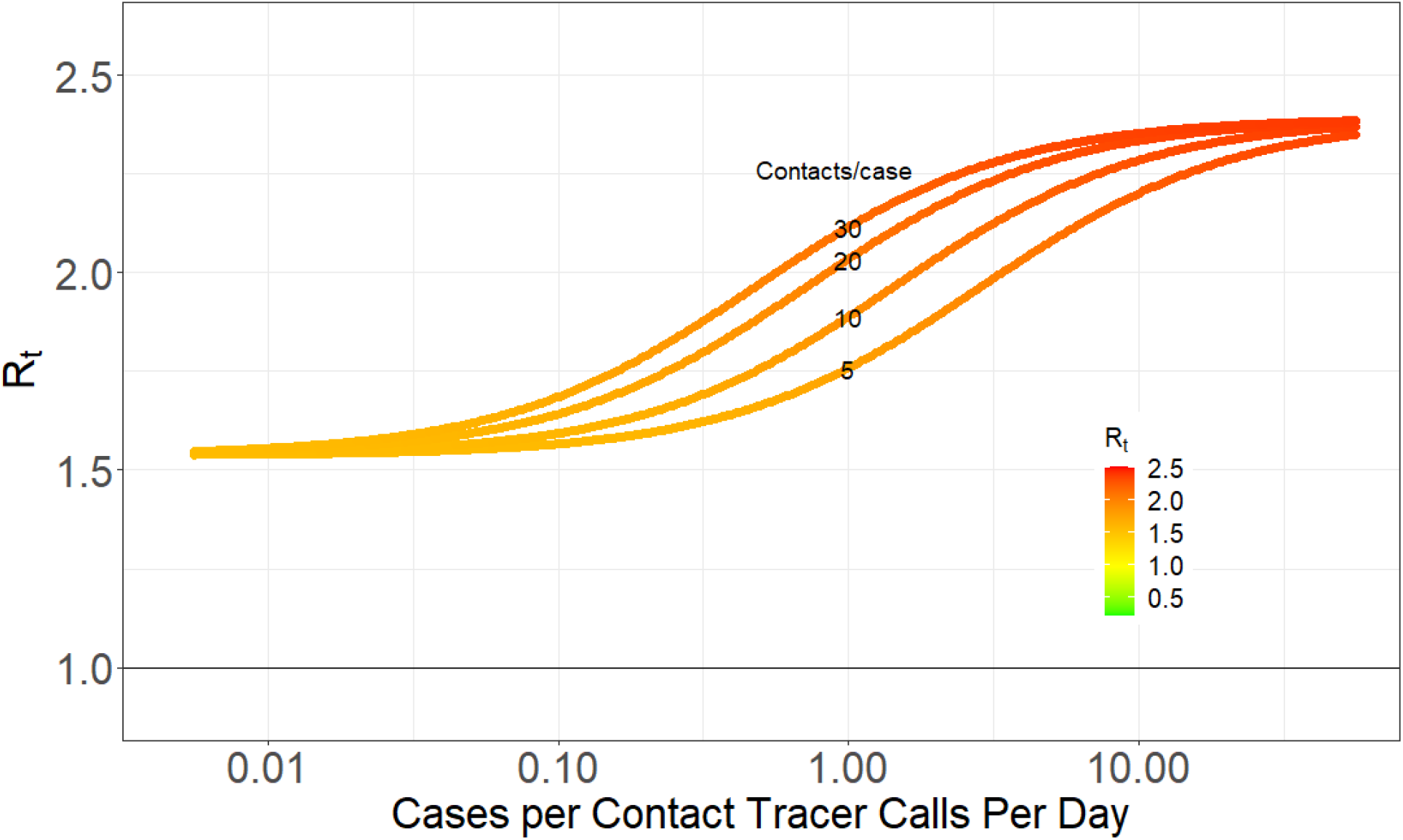
**Pathogen reproductive ratio R_t_ plotted against the number of Cases per contact tracer calls per day for four different number of contacts per case (5, 10, 20, 30; indicated by the small numbers on each curve in the middle of the plot), and an average delay** 1/τ_ms_ **of 5 days between symptom onset and contact tracing (including getting tested and receiving result). With no contact tracing R_t_ = 2.39**.

**Figure 3.**
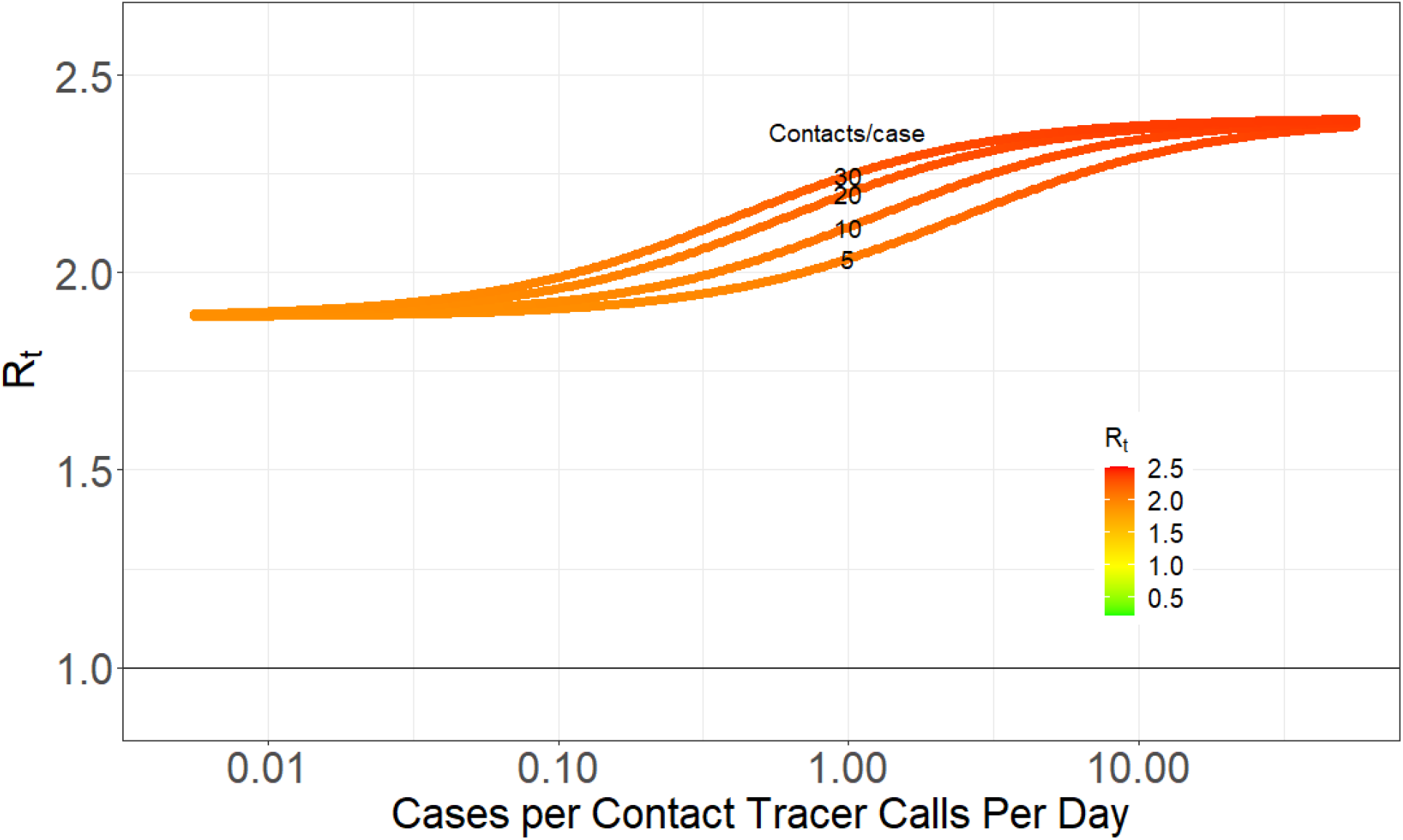
**Pathogen reproductive ratio R_t_ plotted against the number of Cases per contact tracer calls per day for four different number of contacts per case (5, 10, 20, 30; indicated by the small numbers on each curve in the middle of the plot), 12 calls per contact tracer per day, an average delay** 1/τ_ms_ **of 5 days between symptom onset and contact tracing (including getting tested and receiving result). This Figure is the same Figure 2, except only half as many contacts are traced (f_tmstr_ = 0.5). With no contact tracing, R_t_ = 2.39**.

First, the relationship between R_0_ and the number of cases per contact tracer calls per day was sigmoid (Figs 1-3); at both high and low numbers adding or removing contact tracers had smaller effects whereas at intermediate case numbers relative to capacity shifting contact tracers would have a larger impact. When the number of new daily cases per contact tracer call per day was high (i.e. >1 or 10 contacts per contact tracer calls per day), contact tracing had relatively little effect in reducing R_0_ no matter how long the delay was between symptom onset and the start of tracing, 1/τ_ms_ (right side of Figs 1; also evident in Figs 2, 3). This is because contact tracing calls took too long, on average (>10 days), to reduce the infectious period of contacts. Note that for the parameter estimates used (Table 1), an average case becomes infectious starting 3.2 days after infection, and is highly or moderately infectious for an average of 7.3 more days (σ_ps_ and σ_ms_; Table 1). The exponentially distributed durations for the latent and infectious periods implied by standard compartmental models result in only 37% of individuals leaving each class after the average duration, which results in a relatively smaller benefit of contact tracing beyond this case burden, and resultant delay in reaching contacts of cases.

Contact tracers in regions with very high new case numbers relative to contact tracing capacity (>1 on Fig. 1) would have a larger reduction on R_t_ if they were tracing calls with intermediate numbers of cases (0.02 to 1 cases/contact tracer calls per day). However, it is worth noting that reducing R_t_ is not the same as preventing new cases (see Discussion below). Nonetheless, if the goal is to reduce R_t_<1 to stop the growth in cases, the analyses in Figs 1-3 suggest that when new case burdens are high relative to capacity, population-wide interventions (e.g. social distancing or different levels of shelter-in-place orders which reduce contact rates, β or κ, and shift the entire curves in Fig 1-3 downward proportionately; Fig. S2), or orders of magnitude increases in contact tracing capacity will be needed until R_t_ can be effectively reduced by contact tracing.

When there are few (<~0.02) cases per contact tracer call per day, contact tracing was as effective as it could be but excess capacity had little impact, especially for realistic delays, 1/τ_ms_, between symptom onset and the start of tracing (e.g. 3-5 days). Shifting contact tracers to other areas with more cases per contact tracer (0.02 to 0.2) would have minimal impact in increasing Rt but could help substantially reduce Rt in those places with higher new case burdens relative to capacity.

Second, increasing delays, 1/τ_ms_, between symptom onset and the start of tracing greatly influenced the efficacy of contact tracing (Fig. 1). With a 5 day delay, contact tracing could only reduce R_0_ by 33% from 2.6 to 1.6 regardless of contact tracing capacity per case. In contrast, if all symptomatic people sought care and got tested within 1 day of after symptom onset and results were returned within the next 24 hours (1/τ_ms_ = 2 days), contact tracing could reduce R_0_ by 73% from 2.2 to 0.6. Note that the effect of delays in increasing R_0_ due to delayed removal by testing alone (xms) is small compared to the effect of delays in reducing contact tracing efficiency (Fig 1: compare differences among curves in upper right of graph where contact tracing is having little effect to differences among curves in lower left of graph where it has maximal effect). The number of contacts per case obviously also influences the time required to trace these contacts (Fig. 2). If allowable (or illegal) gathering sizes increase, this leads to more contacts per case and a faster decrease in contact tracing efficacy.

Thirdly, if contacts for a substantial fraction of all symptomatic cases do not get traced and quarantined, T-CT-I/Q is far less effective. Figures 1 and 2 showed an optimistic scenario where the fraction of symptomatic infections that are tested and traced is determined only by the delay between symptom onset and testing results being returned (1/τ_ms_) (i.e. all symptomatic infections could be tested and traced). With a 5 day delay (1/τ_ms_=5) (Fig. 2), this results in 38% of infections being detected by testing in the mildly symptomatic state (I_ms_) which is much higher than estimates of case under-ascertainment based on seroprevalence studies (25). If only half of these cases are contact traced (f_tmstr_ = 0.5) the maximum impact of contact tracing is smaller: a 21 % reduction in R_0_ from 2.39 to 1.89 (Fig 3) rather than a 35% decrease from 2.38 to 1.54 (Fig 2).

Reduced contact tracing efficiency with increasing cases can result in a transient accelerating epidemic where R_t_ increases over time (Fig 4). If contact tracing capacity is insufficient to quickly trace contacts, then a decrease in contact tracing efficiency can initially outweigh the depletion of susceptible individuals and lead to an increase in R_t_ over time until depletion of susceptibles overwhelms this effect (Fig 4: compare rightmost panels with limited contact tracing to leftmost panels where social distancing reduces R_t_ to the same initial value as contact tracing). With unlimited contact tracing this phenomenon does not arise (Fig 4: compare middle panels with unlimited contact tracing to leftmost panels).

**Figure 4.**
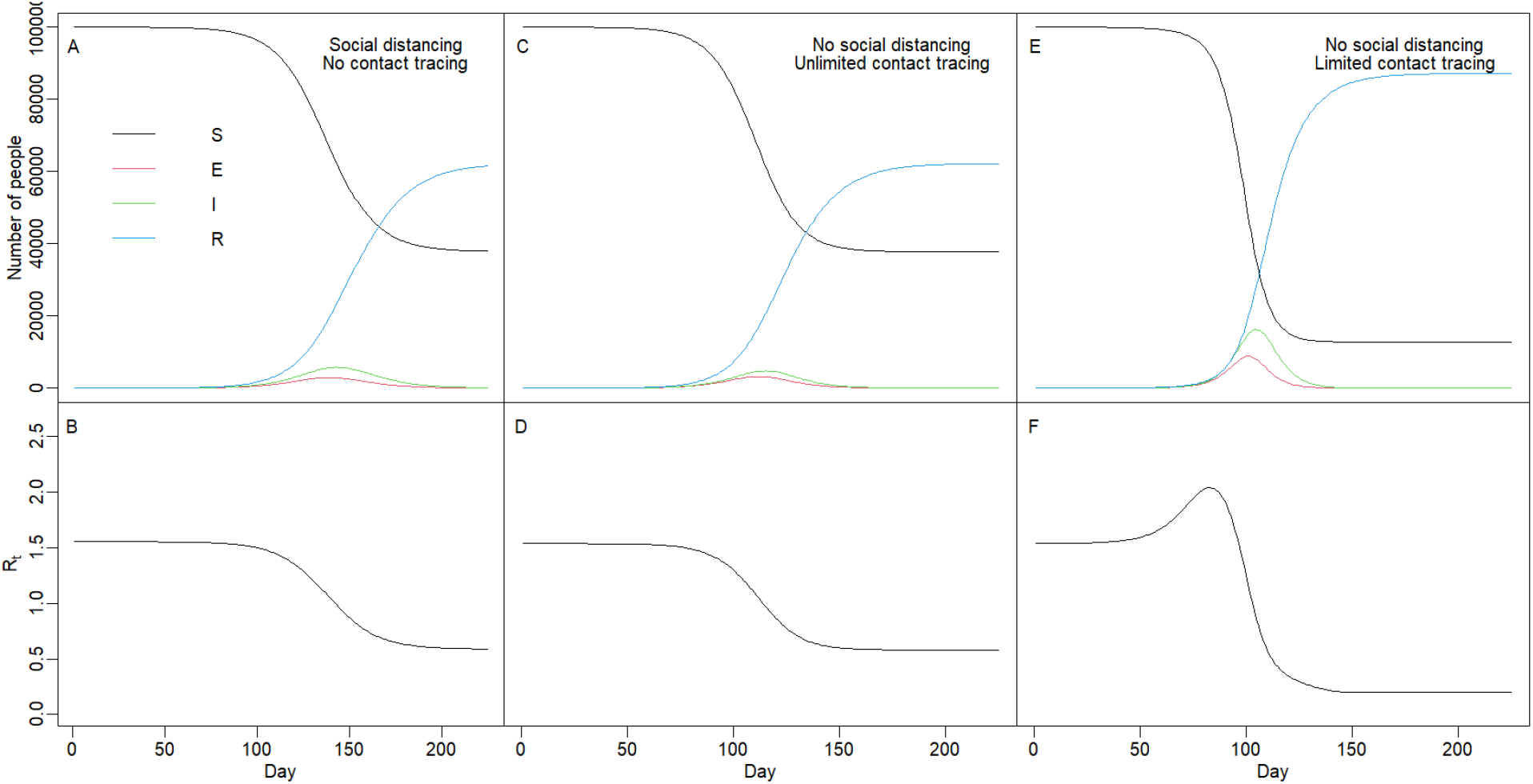
**Impact of reduced contact tracing efficiency with increasing cases. Left panels show (A) the number of susceptible, exposed, infected, and recovered individuals and (B) reproductive number, R_t_, over time with no contact tracing but social distancing (κ = 0.65) set to give same initial R_0_ (1.55) as with contact tracing. Middle panels (C, D) show the same variables with unlimited contact tracing (1500 contact tracers making 12 calls/day; 15 contacts per case;** 1/τ_ms_ **= 5d; f_tmstr_ = 1) but no social distancing (κ = 1). Right panels (E, F) show the same variables and parameters as (C, D) but with <u>limited</u> contact tracing (5 contact tracers)**.

The impact of contact tracing in reducing the pathogen reproductive number R_t_ has two consequences on the temporal timing and establishment of epidemics. First, as is well known, reducing Rt delays and reduces the peak of the epidemic (Fig 5 top vs bottom panels). Second, and less appreciated, stochastic variation in R_t_ can lead to very different timing of epidemics if the initial number of infected individuals is low (Fig 5 left panels), and variation is larger if R_t_ is lower (Fig 5A vs Fig 5C). Finally, heterogeneity in individual transmission can result in local fadeout of the pathogen and this is more if contact tracing reduces R_t_ so that it is closer to 1, and if the number of infected individuals is lower.

**Figure 5.**
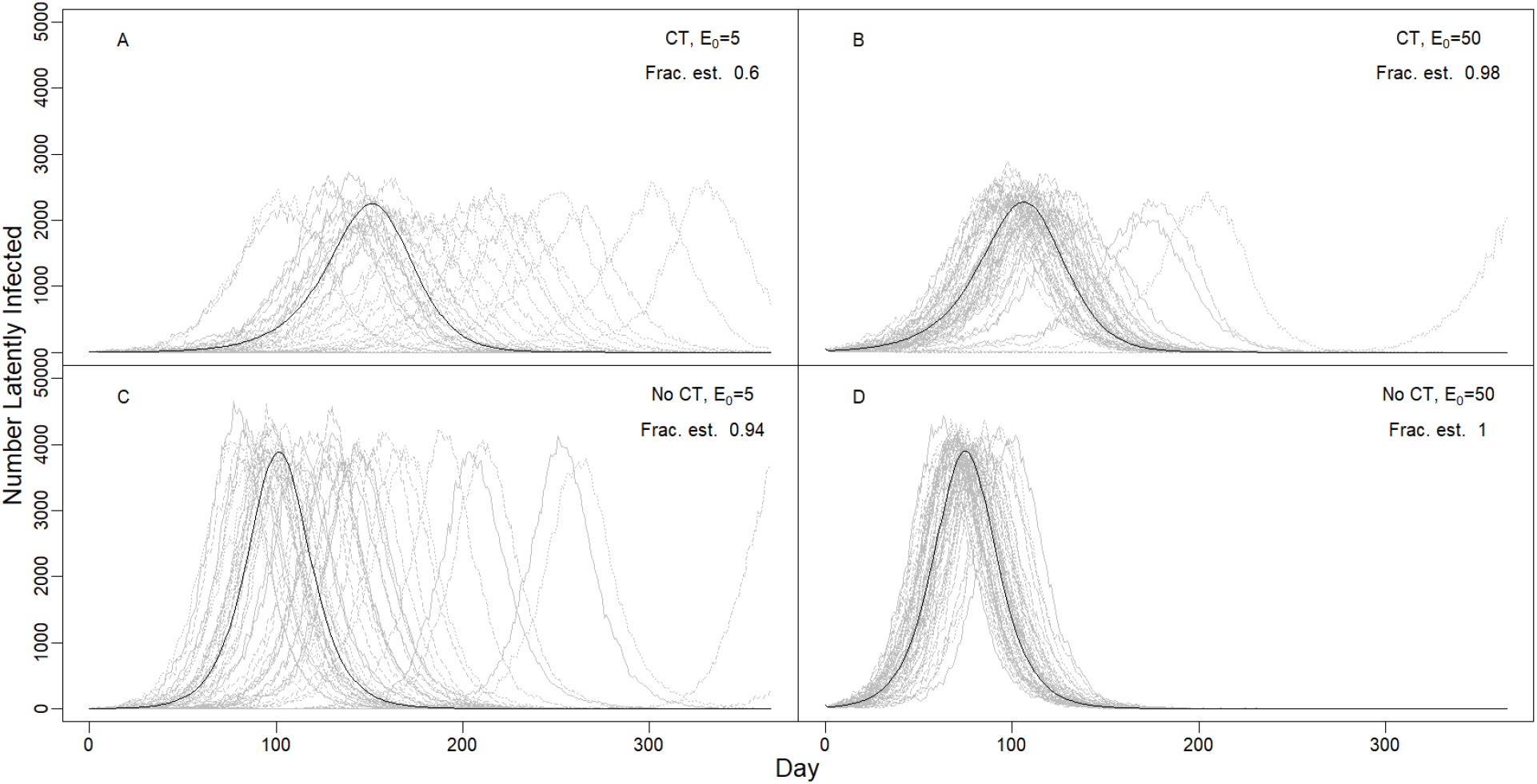
**Variability in the timing and outcome of epidemics due to stochastic variation in individual R_t_. Lines show number of latently infected individuals (in the E class) over time for 1 year with moderate social distancing that reduces contact rates by 30% (κ = 0.7). Grey lines show a single stochastic simulation and the black line shows the deterministic outcome. Frac. est. is the fraction of epidemics that establish (i.e. the maximum number of infected at any time exceeds the starting number infected). Four scenarios include different starting numbers of latently infected individuals on day 0, E_0_ (A, C: 5; B, D: 50), and with (A, B) or without (C, D) contact tracing (CT) which lowered R_0_ from 1.67 to 1.32. The delay from symptom onset to testing and tracing was 5d, but only half of cases were traced (f_tmstr_ = 0.5), as in Fig 3. The modeled population of 100,000 people had 15 tracers making 12 calls/day, and each case had an average of 15 contacts which is intermediate between pre-lockdown and lockdown conditions**.

## Discussion

The two main strategies that have been used to control COVID-19 are T-CT-I/Q and society-wide social distancing interventions (including closing businesses, banning gatherings, wearing masks, etc.) (9). Closing businesses has had devastating impacts on employment and economies, as well as cascading impacts on society. T-CT-I/Q has far smaller economic and societal costs, but its efficacy in controlling epidemics is not fully understood, and some studies suggest that it is insufficient to keep R_t_ below 1 in many settings, especially without digital contact tracing (10, 11, 18). We examined how the efficacy of contact tracing decreases with increasing case burden. At high case burdens relative to contact tracing capacity, contact tracing reached most contacts too late and had little effect on R_t_. Conversely when case numbers were very low relative to contact tracing capacity, there was excess capacity and, all else being equal, contact tracers could be used more effectively in higher case burden settings with very small impacts on local transmission. We note that the exact number of contact tracers needed to reduce R_t_ depends on the number of contacts per case and the number of calls each tracer can make each day (Fig 2). However, the key quantity appears as a ratio of case-contacts per contact tracer calls per day. Thus, each contact tracing team (e.g. a US County) can use local estimates of contacts per case and the number of calls each tracer can make each day to determine where they are on the modelled relationships (Figs 1-3).

A major caveat must be kept in mind in interpreting these results. A smaller reduction in R_t_ (e.g. 10%) in one population can prevent more infections (especially over multiple generations of transmission) than a larger (e.g. 20%) reduction in R_t_ in a second population if R_t_ in the second location is lower (especially when R_t_>1 in the first population), or when there is a larger number of infected individuals in the first population. Thus, transferring contact tracers from a region with a high case burden relative to contact tracing capacity to maximize their efficacy in reducing R_t_ should only be done if other measures (e.g. social distancing) will be put into place to reduce R_t_ where case numbers are high. More generally, allocation of contact tracers to maximize the number of cases prevented given an array of tools would require a complex dynamic analysis beyond that examined here.

We also found that the efficacy of contact tracing itself, regardless of capacity, was strongly influenced by delays between the onset of symptoms and the beginning of tracing, as well as the fraction of symptomatic infections that were traced. Unless delays were short and the fraction of symptomatic cases that were traced was high, contact tracing had limited effects in reducing R_t_. This finding parallels results from other studies demonstrating the large effects of delays in reducing efficacy of isolating infections by testing alone (26). We note that in the model considered here, only symptomatic individuals were removed by testing (pre-symptomatic and asymptomatic infections were not tested) which leads to a much smaller impact of testing on R_t_. Our results emphasize the importance of encouraging people to get tested as soon as possible after mild symptom onset, and having sufficient testing capacity to return their results quickly. Similarly, the fraction of symptomatic infections that get tested and traced is poorly known, but if the ratio of infections to cases from seroprevalence studies in some locations is approximately correct (e.g. 10:1; (25)), then contact tracing will have very limited effects in reducing transmission.

Allocation of contact tracing resources can be most efficiently deployed in two ways. First, contact tracing is much more effective when infections are detected soon after symptom onset. One should prioritize these individual cases for tracing since their contacts are likely to be earlier in their infections and quarantining them will cut off most or all of their infectious period. If one knows the date of contact between the case and the contact, one could also prioritize tracing more recent contacts and those that had contact with the case during the case’s days of peak infectiousness just before and after symptom onset (27, 28). Second, if one is attempting to limit transmission in multiple regions (e.g. US counties within a state) one could deploy contact tracers to counties where they will be able to have the most impact: from places with excess capacity to those with intermediate numbers of cases per contact tracer calls per day. Conversely, if contact tracers cannot quarantine the contacts of cases within 10-12 days of the case’s symptom onset, they will be unlikely to effectively reduce transmission from those contacts.

Our results also offer insight on two phenomena observed in COVID-19 epidemics that are not fully understood. First, epidemic dynamics sometimes differ enormously between places that seem otherwise similar. This may be due to differences in social behavior or contact patterns, but our results illustrate that stochastic chance may also play a role in shifting the timing of epidemics by more than a month, especially when the initial number of cases is low and R_t_ is closer to 1 (i.e. when lockdowns are first lifted) so that populations spend longer periods of time with few cases where stochastic variation is most important. Second, staged business re-openings have sometimes led to accelerating or runaway epidemics. These may be due to sudden changes in social behavior, but decreases in contact tracing efficiency may also contribute to these accelerating epidemics, if contact tracing was playing a significant role in limiting transmission. Increasing contact tracing capacity could limit this epidemic acceleration as cases increase, which suggests that training a reserve capacity of tracers to be used following case surges or being able to deploy a mobile tracing force could help limit runaway epidemics.

More broadly, contact tracing could play an important role in limiting transmission of SARS-CoV-2 and other pathogens. However, we found that its efficacy depends on participation in seeking testing immediately following symptom onset and quick return of test results, as well as sufficient contact tracing capacity if case numbers surge. Shortcomings in each of these factors greatly limit its efficacy, which could lead to implementation of much more damaging measures to control transmission, including widespread business and school closures. Investments in public health, including testing, contact tracing, and public outreach to encourage health seeking when symptomatic, is likely a much more cost-effective approach to control COVID-19.

## Data Availability

There are no original data in the manuscript. A link to a Github repository with code to replicate the results is provided at the end of the methods.

## Acknowledgements

We thank the dozens of scientists and public health practitioners who have informed this work through comments and discussions.

---

**Figure S1.**
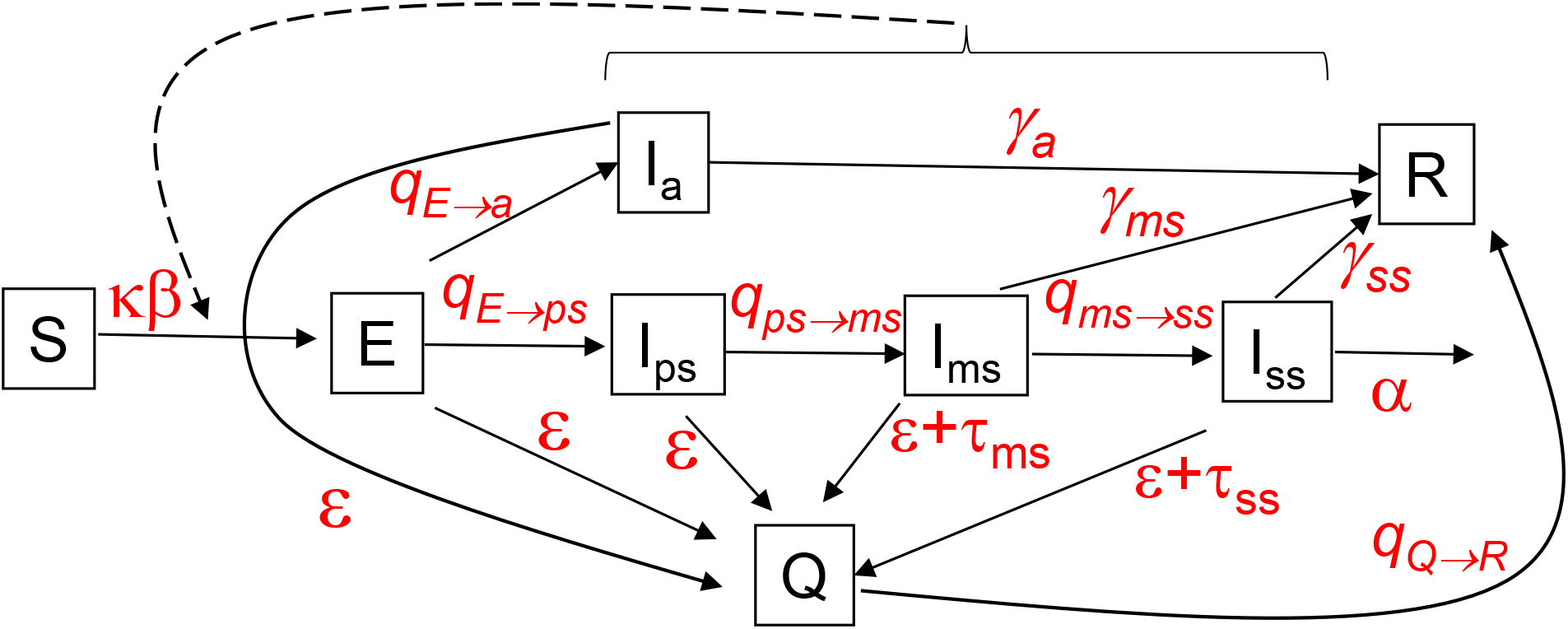
**Compartmental model of SARS-CoV-2. See text for equations and parameter values. Boxes represent Susceptible (S), Exposed (E), Infected (I), recovered (R), and Quarantined (Q) classes. There are four compartments for infected individuals that reflect symptom severity (asymptomatic**, ***I_a_,* pre-symptomatic**, ***I_ps_,* mildly symptomatic**, ***I_a_,* and severely symptomatic**, ***I_ss_*) . κ is a social distancing factor between 0 and 1 that modifies the contact rate β, σ are infectiousness for each of the I classes, q are transition rates between classes given by the subscripts separated by the arrow (q_E→ps_ is the transition rate between the E and I_ps_ classes), ε is the rate of removal by contact tracing from the E or I classes to the quarantined class Q, τ are the removal rate by testing of mildly symptomatic or severely symptomatic infected individuals, α is the disease-caused death rate, and γ are the recovery rates to the R class. The dashed line and bracket indicate that all 4 classes of infected individuals contribute to transmission**.

**Figure S2.**
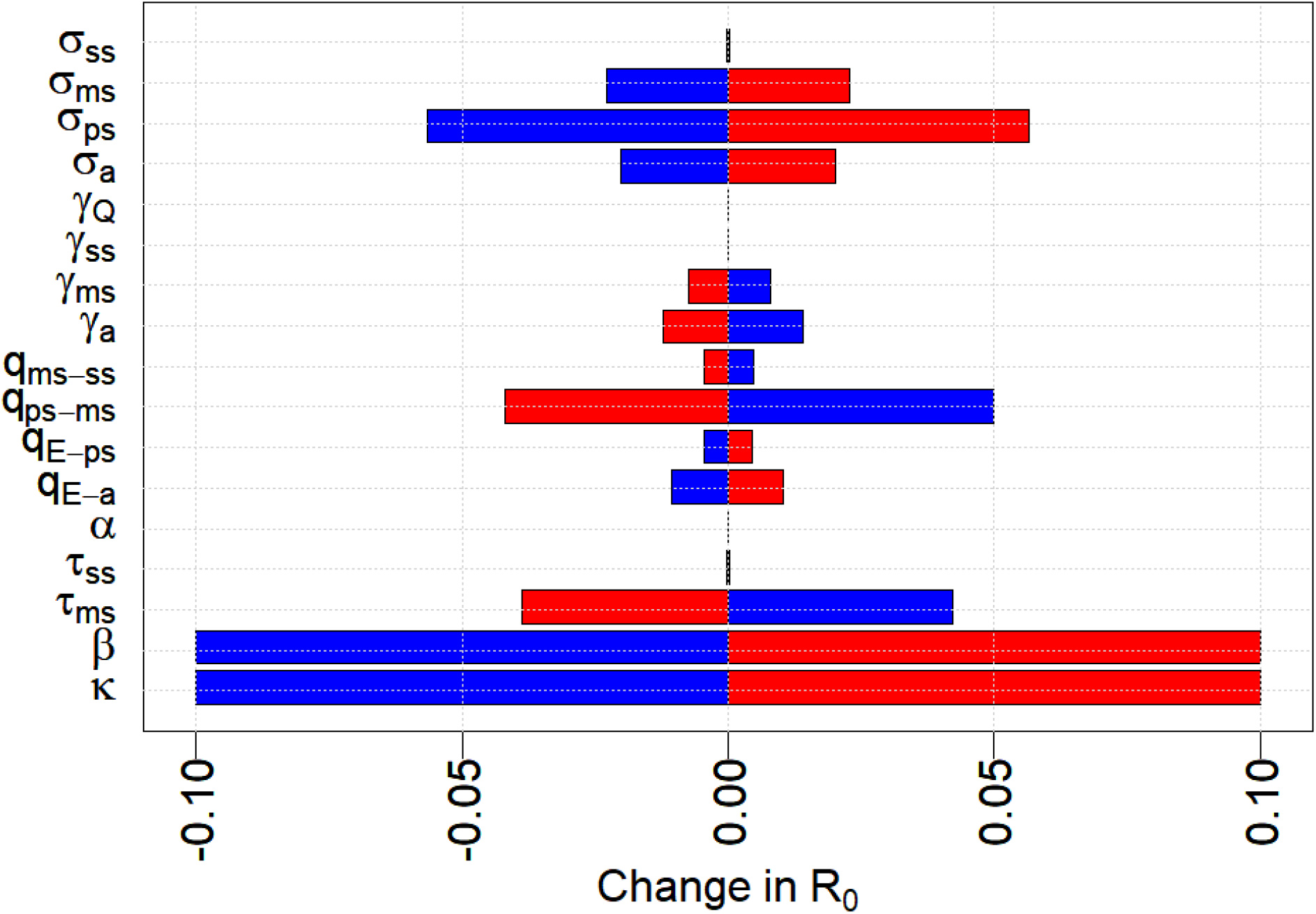
**Sensitivity analysis. The plot shows how much R_0_ changes from a ten percent increase (red) or ten percent decrease (blue) in that model parameter relative to values in Table 1 (with τ_ms_ = 0.2; f_tmstr_ = 1; κ = 1). R_t_ scales linearly with β and κ, whereas q_ps→ms_, τ_ms_, and σ_ps_ have half as large an effect as β or κ and other parameters are even less influential**.

